# High rates of diabetes and pre-diabetes in postpartum period in Indian GDM women

**DOI:** 10.1101/2021.01.04.21249193

**Authors:** S Yajnik Chittaranjan, S Wagle Sonali, R Shukla Sharvari, D Kale Shailaja, S Ladkat Rasika, G Lubree Himangi, S Bhat Dattatray, S Memane Nilam, Sanat Phatak, K Meenakumari

**Author notes:** Corresponding author: Prof Chittaranjan S. Yajnik., Director, Kamalnayan Bajaj Diabetology Research Centre, Diabetes Unit, King Edward Memorial Hospital Research Centre, Rasta Peth, Pune, 411011, India. **Presentation at a meeting**: No.

## Abstract

**Aim:** To investigate postpartum glucose intolerance in South Asian Indian GDM women within 1 year of delivery.

**Methods:** Between 2001-2005, 220 women were treated for GDM at the Diabetes Unit, King Edward Memorial Hospital, Pune. GDM was diagnosed by 75g OGTT [WHO 1999 pregnancy criteria]. OGTT was repeated 3 months to 1 year postpartum. One hundred and nineteen non-GDM women were also studied.

**Results:** Of 220 GDM women [30years, BMI 26.0kg/m^2^] 9 women continued to be diabetic after delivery and a further 111 attended OGTT within one year of delivery. Two had IFG, 16 IGT and 23 diabetes [WHO 1999], thus 50[42%] women were glucose intolerant. Of the non-GDM, 1 had IFG, 8 IGT and 3 diabetes (10% glucose intolerant). Those who were hyperglycemic at follow up had stronger family history of diabetes [64% vs. 58%], were shorter [154.6 vs. 156.5cm], had higher FPG concentrations during pregnancy [5.27 vs. 4.99 mmol/L], and higher BMI [26.3 vs.25.0kg/m^2^] and waist circumference [88.0 vs. 82.3cm] at follow up compared to normoglycemic women. Hyperglycemia was not associated with GAD antibody positivity (4 vs 3 in normoglycemic).

**Conclusion:** We describe one of the highest rates of postpartum hyperglycemia within a short time after delivery in young urban GDM women from India. Majority of risk factors for GDM were present from before pregnancy, and we propose that metabolic disturbances were also likely present. This has implications for peri-conceptional epigenetic programming of diabetes in the offspring. Pre-pregnancy screening and treatment of glucose intolerance and its risk factors in the high-risk populations could be an important measure for primordial prevention of diabetes.

**Key messages:**

- We describe one of the highest rates of postpartum hyperglycemia in young urban GDM women from India within a short time after delivery.
- Our results invite further research and policy discussion for screening and treatment of glucose intolerance before pregnancy in high-risk populations.

## Introduction

The classic study by O’Sullivan’s group in Boston, USA showed that∼50% of the women diagnosed with gestational ‘hyperglycemia’ developed glucose intolerance in subsequent years. ^*[1]*^ This became the basis for description of the entity ‘Gestational diabetes mellitus’ [GDM]. It was soon recognized that pregnancy hyperglycemia affected not only maternal but also fetal outcomes, both in short and long term. Thus GDM achieved the status of a high-risk pregnancy as well as being a window on the future health of the mother and the baby.

The current definition of GDM: “any level of glucose intolerance first recognized during pregnancy” ^*[2]*^ is insensitive to the contribution of undiagnosed pre-gestational glucose intolerance or to its continuation postpartum. There is little systematic study of contribution of pre-pregnancy glucose intolerance to ‘GDM’, though a substantial proportion of risk factors for GDM are present before conception. ^*[3, 4]*^

On the other hand, there are many studies of post-partum follow up of GDM women. A recent review showed that 2.6% to 70% women with GDM in the low risk populations are diabetic when examined 6 weeks to 28 years postpartum, respectively. ^*[5]*^ In Chennai (India), 20% of GDM women diagnosed by universal screening using IADPSG criteria were glucose intolerant within one year after delivery: 12% within 6-12 weeks after delivery. ^*[6]*^ A follow up of GDM women in Pune showed that, 52% had diabetes and 15% IGT at an average of 4.5 years after delivery. ^*[7]*^ A tacit assumption in all these reports is that GDM is a transient condition: they have a ‘normal’ glucose tolerance before conception, become glucose intolerant during pregnancy, and become ‘normal glucose tolerant’ after delivery. The subsequent diabetes is considered a ‘new’ entity. In clinical practice, high glucose levels in early pregnancy or persistence of glucose intolerance after delivery is interpreted as indicative of ‘pre-gestational’ diabetes. In populations with high prevalence of diabetes in the young, it is possible that a sizable proportion of ‘GDM’ could be undiagnosed pre-gestational diabetes discovered in pregnancy.

Given the high rates of diabetes and pre-diabetes within five years of delivery in our GDM women, we decided to investigate the risk factors for glucose intolerance within one year after delivery. This could be a pointer towards pre-gestational risk of diabetes, and could advise preconceptional screening for risk factors and glucose intolerance in young Indian women.

## Material and methods

Between 2001 and 2005, we treated 220 women for GDM at the Diabetes Unit, King Edward Memorial Hospital, Pune. Obstetricians in our hospital prescribed an oral glucose tolerance test, [OGTT] usually to high-risk pregnant women [family history of diabetes, history of GDM, having delivered a large baby, bad obstetric history, overweight and obesity, excessive weight gain during pregnancy, glycosuria, hypertension, polyhydramnios and/or macrosomia on ultrasonography]. Diagnosis of GDM was based on WHO 1999 pregnancy criteria (75g anhydrous glucose OGTT). ^*[8]*^ All women diagnosed with GDM were advised on diet and physical activity. If blood glucose concentrations were still elevated after 7 days of lifestyle adjustment [Fasting ≥ 5.0 mmol/L and 2-hour post meal ≥ 6.6mmol/L], anti-diabetic medications [insulin or oral anti diabetic agents [OADs]] were prescribed and home monitoring of blood glucose advised. Doses were adjusted to achieve the targets. At delivery, following data was obtained from hospital records: medical and obstetric history, the time and results of glucose tolerance test and the time and mode of delivery. At discharge from the hospital, anti-diabetic treatment was continued if glucose concentrations were: fasting >6.1mmol/L, post-meal >7.7mmol/L; if below these, they were instructed to attend an oral glucose tolerance test [OGTT] after three months.

At the time of follow-up [3 months to 1 year postpartum], an OGTT was performed and anthropometry, blood pressure and biochemical parameters were measured. Height was measured to the nearest 0.1cm using a wall fixed stadiometer [CMS Instruments, London, UK] and body weight was recorded to the nearest 0.1 kg using a digital weighting scale [Soehnle, Waagen, GmBH, Germany]. The biceps, triceps, subscapular and suprailiac skinfold thicknesses were measured to the nearest 0.2 mm on non-dominant side of the body using Harpenden skinfold calipers[CMS Instruments, London, UK]. Waist circumference was measured to the nearest 0.1 cm using a non-stretchable fiberglass measuring tape[CMS Instruments, London, UK]. Two blood pressure readings were recorded in supine position after 5 minutes rest with a digital monitor [UA 767PC, A & D Instruments Ltd., Abingdon, UK]. Mean of two measurements was used in analysis.

Venous blood samples were obtained fasting and 2 hours after glucose drink. Following measurements were made using standard enzymatic kits: plasma glucose, total cholesterol, HDL-cholesterol, triacylglycerol and insulin. We used WHO 1999 non-pregnant criteria for classification of glucose tolerance [8]. The glutamic acid decarboxylase (GAD) antibody was measured using Auto DELPHIA Kit [PerkinElmer, Turku, Finland].

We also followed women [age and socioeconomically matched from hospital records] who had uncomplicated full term pregnancies during the same period and whose fasting plasma glucose concentrations were ≤ 5.0mmol/L and 2-hour post meal ≤ 6.6mmol/L during pregnancy. These are considered as ‘non-GDM controls’. They underwent similar measurements.

### Definitions

1. Diagnosis of GDM was based on WHO 1999 Pregnancy criteria [fasting plasma glucose ≥ 6.1mmol/L and 2-hour plasma glucose ≥ 7.7 mmol/L] using a 75 g OGTT.
2. In non-pregnant state, diagnosis of DM, IFG and IGT was based on WHO 1999criteria[DM: fasting plasma glucose ≥7.0mmol/L or2-hour plasma glucose ≥11.0mmol/L IGT: 2-hour plasma glucose ≥7.7 mmol/L, IFG: fasting plasma glucose ≥6.1mmol/L and <7.0mmol/L].
3. Insulin resistance and beta cell function [HOMA-IR and HOMA-β respectively] were calculated using the online Oxford HOMA Calculator [http://www.dtu.ox.ac.uk].
4. Disposition index was calculated as HOMA-β/HOMA-IR.
5. GAD positive was defined as GAD antibody value ≥12.8 unit[As per the Kit].
6. Metabolic syndrome was defined as per International Diabetes Federation [IDF] 2006 criteria. ^*[9]*^
7. Gestational hypertension or pregnancy-induced hypertension (PIH) is the development of new hypertension in a pregnant woman after 20 weeks gestation without the presence of protein in the urine or other signs of preeclampsia. Hypertension is defined as having a blood pressure greater than 140/90 mm Hg.

### Statistical methods

Data are presented as mean (SD) and n[%]. For statistical analysis, variables with skewed distributions were log-transformed to satisfy assumptions of normality. The mean difference between the groups was analyzed using ANOVA. We performed logistic regression analyses to study the factors predicting hyperglycemia at the time of follow up. Analysis was carried out using Statistical Package for Social Sciences [SPSS] 16.0 version.

### Ethics

Informed consent was obtained from the participants and the KEM Hospital Research Centre’s Ethics Committee approved the study.

## Results

Of 220 GDM women, 46% responded adequately to lifestyle adjustment, 4.0% were treated with OADs and 50% with insulin. Seven percent women developed pregnancy-induced hypertension [PIH], 53% were delivered by Cesarean section, and 4% neonates were admitted to NICU for treatment.

### Follow up

Nine women were continued on anti-diabetic treatment at discharge from the hospital because of persistent hyperglycemia. We were able to review additional 111 women within one year of delivery [72% within 6 months]. These women did not differ significantly from those who did not respond to our invitation for postpartum follow up [n=100] with respect to standard risk factors for diabetes [age, family history of diabetes, BMI] and level of glycemia at diagnosis of GDM [p>0.05 for all, data not shown]. We were also able to study 119 mothers who were not diagnosed with GDM [non-GDM controls].

Of 111 GDM women, two had impaired fasting glucose [IFG], 16 impaired glucose tolerance [IGT] and 23, diabetes [T2D]. Thus 50 of 120 [42%] women were glucose intolerant within 12 months of delivery, 32 [27%] being diabetic. Of the 119 non-GDM women, one woman had IFG, eight IGT and three DM i.e. 12 of 119 [10%] were glucose intolerant, only three [2%] being diabetic **[Figure 1]**.Women who were hyperglycemic at follow up [from both the GDM and non-GDM groups] had higher BMI, waist circumference, body fat percent, triacylglycerol concentrations, HOMA R, lower disposition index and higher blood pressure compared to normoglycemic women. Irrespective of their glucose tolerance status in pregnancy, these women had delivered heavier babies and had a higher rate of Cesarean section **[Table 1]**.

**Table 1:** Anthropometric and biochemical characteristics of Non-GDM control and GDM women at review [within 1 year after delivery] according to current glycemic status [by World Health Organization 1999 non pregnant criteria].

|  | Non GDM control |  |  |  | GDM |  |  |  |  |  |  |
| --- | --- | --- | --- | --- | --- | --- | --- | --- | --- | --- | --- |
|  | n | Normoglycemic | n | Hyperglycemic | n | Normoglycemic | n | Hyperglycemic | p1 | p2 | P3 |
| **N** |  | 107 |  | 12 |  | 70 |  | 50 |  |  |  |
| **During pregnancy** |  |  |  |  |  |  |  |  |  |  |  |
| Fasting glucose [mmol/L] | 56 | 4.27[0.44]<sup>*</sup> | 9 | 4.27 [0.49] | 58 | 5.72[1.55] | 34 | 6.21[1.67] | 0.0001 | 0.217 | 0.079 |
| 2 hour glucose [mmol/L] | 71 | 5.49[0.88]<sup>*</sup> | 8 | 5.60[0.72] | 56 | 9.26[2.38] | 37 | 9.99[2.78] | 0.0001 | 0.109 | 0.019 |
| Gestation at diagnosis [weeks] |  | NA |  | NA | 49 | 29.0 [6.5] | 35 | 26.6 [7.7] | -- | 0.366 | -- |
| **At delivery** |  |  |  |  |  |  |  |  |  |  |  |
| Cesarean section | 44 | 44 [41.2] | 10 | 10 [83.3] | 65 | 37 [52.2] | 40 | 36 [72.5] | 0.043 | 0.030 | 0.049 |
| Birth weight [Kg] | 78 | 2.83 [0.42] | 5 | 3.08 [0.44] | 53 | 2.82 [0.48] | 27 | 2.91 [0.46] | 0.036 | 0.907 | 0.078 |
| **At Review** |  |  |  |  |  |  |  |  |  |  |  |
| Duration since delivery [years] | 103 | 0.45 [0.23] | 12 | 0.38 [0.19] | 70 | 0.48 [0.20] | 50 | 0.46 [0.19] | -- | -- | -- |
| Age [years] | 103 | 28.2 [3.5] | 12 | 29.9 [4.7] | 62 | 28.5 [4.4] | 44 | 31.9 [3.4] | -- | -- | -- |
| Height [cm] | 103 | 156.5 [5.1]<sup>*</sup> | 12 | 156.2 [6.4] | 64 | 154.6 [5.3] | 46 | 156.9 [5.2] | 0.058 | 0.928 | 0.071 |
| Body Mass Index [Kg/m<sup>2</sup>] | 103 | 24.0 [3.8] | 12 | 25.3 [3.4] | 64 | 25.1 [3.7] | 46 | 27.4 [4.8] | 0.001 | 0.093 | 0.020 |
| Waist circumference [cm] | 103 | 80.9 [9.7] | 12 | 86.1 [8.6] |  | 83.7 [9.6] |  | 89.9 [10.0] | 0.0001 | 0.075 | 0.010 |
| Blood pressure [mmHg] | 103 | 110/68 [10/9] | 12 | 117/70 [10/13] | 64 | 109/69 [13/12] | 46 | 119/76 [13/11] | 0.061 | 0.006 | 0.992/0.736 |
| **Biochemical characteristics** |  |  |  |  |  |  |  |  |  |  |  |
| Hemoglobin [g/dl] | 103 | 12.4 [0.98] | 12 | 12.8 [1.1] | 65 | 12.6 [1.1] | 48 | 12.7 [1.4] | 0.059 | 0.500 | 0.251 |
| Fasting glucose [mmol/L] | 103 | 4.66[0.44]<sup>*</sup> | 12 | 5.45[0.99] | 70 | 4.85[0.49] | 50 | 6.88[2.51] | 0.0001 | 0.0001 | 0.076 |
| 2 hour glucose [mmol/L] | 103 | 5.55[0.94]<sup>*</sup> | 12 | 8.78[2.44] | 70 | 5.85[0.90] | 50 | 11.8[4.86] | 0.0001 | 0.0001 | 0.019 |
| Fasting insulin [pmol/L] | 99 | 45.0[30.6] | 12 | 77.4[83.4] | 68 | 66.0[68.4] | 43 | 78.0[64.2] | 0.0001 | 0.064 | 0.053 |
| HOMA-IR | 99 | 1.08 [0.73] | 12 | 1.95 [2.09] | 68 | 1.57 [1.49] | 43 | 2.06 [1.53] | 0.0001 | 0.012 | 0.015 |
| HOMA-β | 99 | 109.6 [50.9] | 12 | 114.2 [54.05] | 68 | 133.3 [83.1] | 43 | 97.8 [58.5] | 0.0001 | 0.057 | 0.048 |
| Disposition index | 99 | 127.2 [62.3] | 12 | 86.9 [45.7] | 68 | 108.5 [56.0] | 43 | 58.5 [33.4] | 0.0001 | 0.0001 | 0.018 |
| Total cholesterol [mg/dl] | 103 | 4.19[0.83] | 12 | 4.67[1.14] | 66 | 3.97[0.94] | 49 | 4.31[1.03] | 0.294 | 0.094 | 0.098 |
| HDL cholesterol [mg/dl] | 103 | 1.20[0.24]<sup>*</sup> | 12 | 1.37[0.21] | 66 | 1.10[0.26] | 49 | 1.14[0.29] | 0.0001 | 0.495 | 0.002 |
| Triacylglycerol[mg/dl] | 103 | 0.78[0.32] | 12 | 1.10[1.12] | 66 | 0.85[0.36] | 49 | 1.27[0.71] | 0.0001 | 0.001 | 0.125 |
| Metabolic syndrome | 103 | 0 | 12 | 7 [58] | 70 | 0 | 45 | 27 [54] | 0.0001 | -- | -- |

Values are mean [SD] or n[%].
p1- Significance value between Non-GDM control and GDM women, adjusted maternal age and years since delivery to follow up.
p2- Significance value between hyperglycemic and normoglycemic women from GDM group, adjusted maternal age and years since delivery to follow up.
p3- Significant difference between normoglycemic women in Non-GDM control and GDM group, adjusted maternal age and years since delivery to follow up

**Figure 1.**
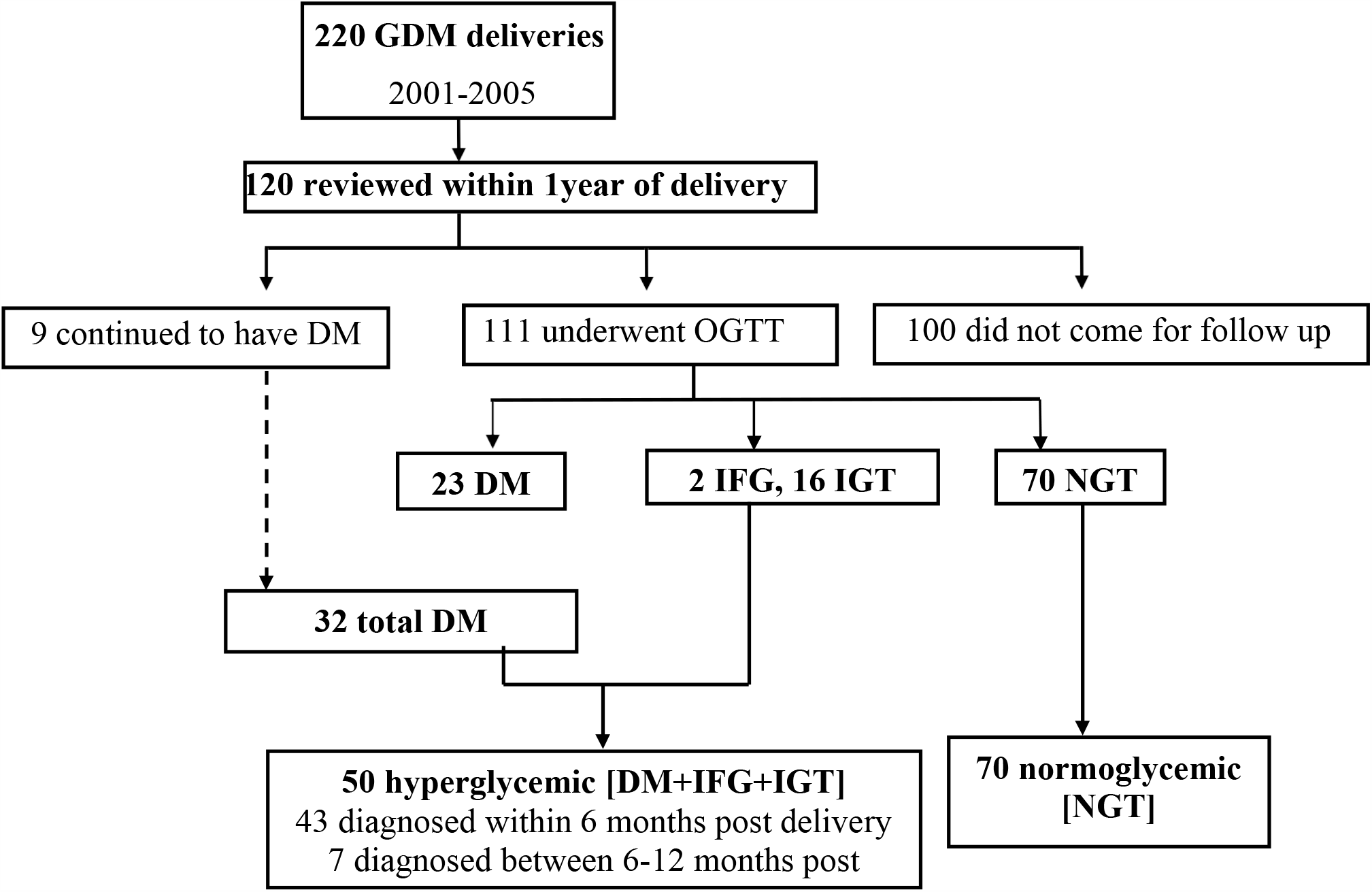
A flow diagram to describe the study subjects and their glycemic status at follow up

On multiple logistic regression, hyperglycemia at follow up was significantly associated with higher maternal age [p<0.05], higher HOMA-R [p<0.01] and lower disposition index at follow up [p<0.05]. At follow up, GDM women (irrespective of their glucose tolerance) were more obese [BMI 26.0 vs. 24.2 kg/m^2^, p<0.001], more centrally obese [waist 86.4 vs. 81.4 cm, p<0.001] and more adipose [sum of skinfolds [59.3 vs 50.5 mm, p<0.01] compared to non-GDM controls. They also ‘had lower plasma HDL [1.12 vs 1.23 mmol/L, p<0.001], higher triacylglycerol concentrations [1.01 vs 0.80mmol/L, p<0.001], higher insulin resistance[HOMA-R 1.76 vs 1.17, p<0.001], lower disposition index[86.5 vs 121.3, p<0.05] and higher blood pressure [114/72 vs. 111/68 mm of Hg, p<0.05]. Twenty-four percent of GDM women and 6% of non-GDM women fulfilled the IDF criteria for metabolic syndrome. GAD antibody was measured in 89 GDM women, and was positive in 7; of these 3 were hyperglycemic and 4 normoglycemic at follow up.

GDM mothers who were normoglycemic at follow up were shorter and had higher glucose and insulin concentrations, higher HOMA-IR, lower disposition index and lower HDL concentrations compared to non-GDM control mothers who remained normoglycemic at follow up.

We found a gradient of increasing levels of risk factors for diabetes (age, obesity and central obesity, higher glucose and compromised beta cell function) starting from women who were NGT-NGT to those who were GDM-hyperglycemic through the intermediate categories **[Figure 2]**. The categories include GDM-Hyperglycemic (n=50), Non-GDM Hyperglycemic (n=12), GDM-Normal (n=70) and Non-GDM-Normal (n=107). The graded distribution is suggestive of an ongoing phenotype across pre-pregnant, pregnant and post-delivery period.

**Figure 2:**
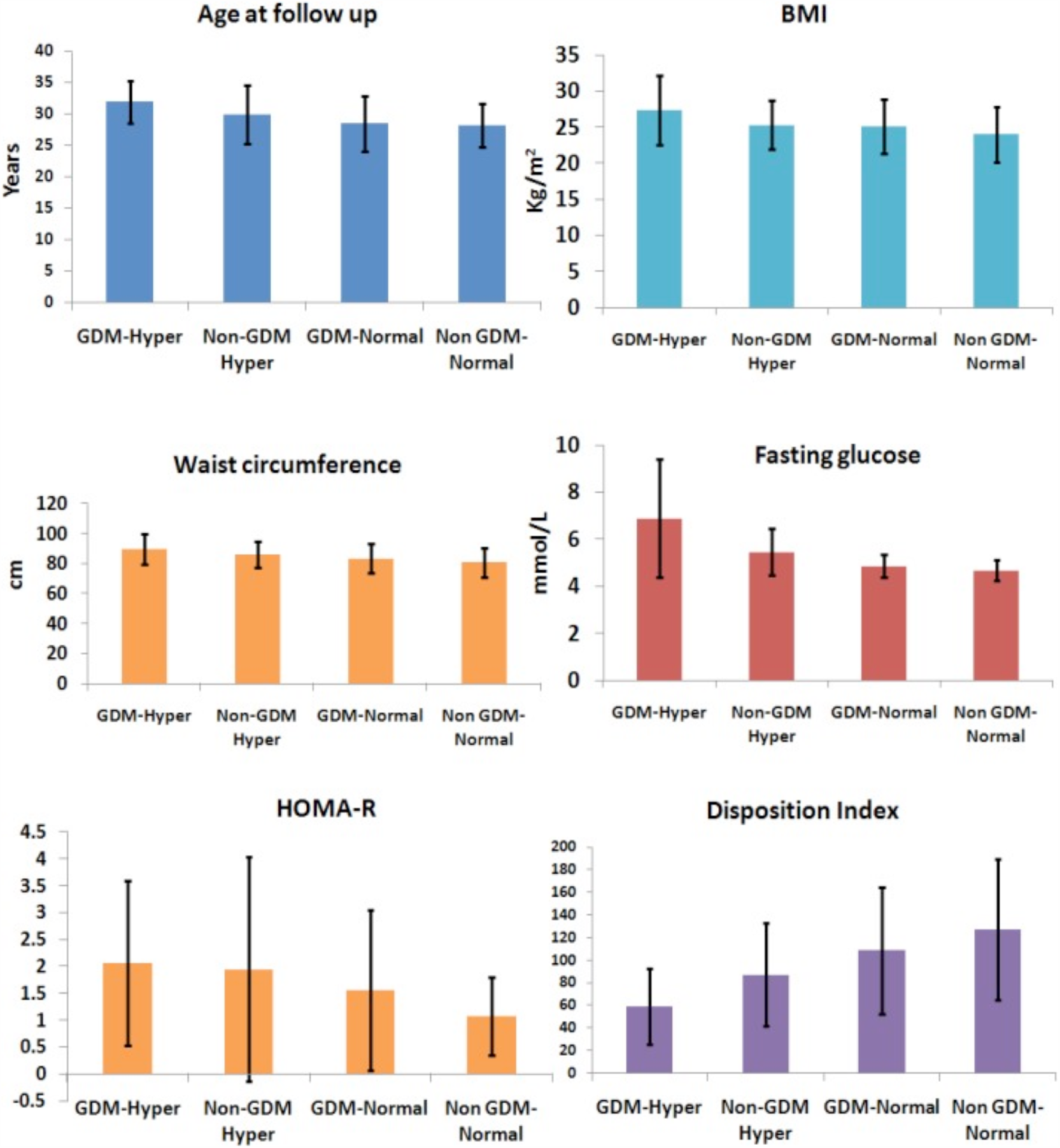
*Analysis of trend was significant for all the measurements The distribution of diabetes risk factors in GDM women and non-GDM women who were hyperglycemic or normoglycemic at follow up*

## Discussion

In a post-partum follow up of high risk GDM women [WHO 1999 Criteria] within one year of delivery, we found 27% had diabetes and a further 15% had pre-diabetes. This is one of the highest reported rates of glucose intolerance in GDM women within a short time of delivery and is suggestive of continuation of pre-gestational glucose intolerance through pregnancy. This conclusion is supported by significantly higher levels of glucose concentrations and other diabetes related risk factors in the ‘normoglycemic’ GDM compared to normoglycemic non-GDM women. There was an increasing gradient of diabetes risk factors: age, BMI and waist circumference, from non GDM-normoglycemic to GDM-hyperglycemic women through the intermediate groups. We feel that this suggests a continuation of an existing phenotype, probably from before pregnancy. Majority of GDM women have T2D characteristics, a few women were GAD antibody positive but none had T1D. Our findings raise a disconcerting possibility that the current guidelines and practice of ‘searching’ for ‘GDM’ in pregnancy may systematically distract us from the opportunity to detect and correct pre-conceptional maternal metabolic and physical risk factors which influence intergenerational epigenetic transmission of susceptibility to obesity, diabetes and other NCDs. ^*[10]*^

Our findings are not entirely surprising, given the high prevalence of glucose intolerance and its risk factors in young Indians. ^*[11-14]*^ Similar findings may be expected in other populations of the developing countries, which contribute 80% to the world’s diabetes population [*www.idf.org/diabetesatlas*]. Average age at diagnosis of diabetes and CVD is 5-10 years younger in South Asian Indians compared to Europeans ^*[15,16]*^, and the prevalence of diabetes in Asian children is 9 times higher than the white children in the UK. ^*[17]*^ The National Urban Diabetes Survey [NUDS] showed a prevalence of 2.4% diabetes and 12.2% pre-diabetes in 20-30 years old Indian women in the year 2000. ^*[11]*^ INDIAB study showed that in urban Maharashtrian women hyperglycemia (pre-diabetes+diabetes) rapidly increases from 25 years of age. ^*[12]*^ At 18 years of age, 18% of women in the rural Pune Maternal Nutrition Study were pre-diabetic based on ADA 2014 criteria. ^*[13]*^ These women had higher glucose and lower disposition index from childhood and early adolescence (6, 12 years of age) [18] Similarly, diabetes at 40 years of age was predicted by higher childhood glucose in the cohorts in the i3C study. [19] A retrospective search of records of women diagnosed with ‘GDM’ in the Kaiser Permanente clinic showed higher glucose and lipid abnormalities years before pregnancy.

The current practice of screening for GDM in the third trimester of pregnancy is based on experience in low prevalence western populations many decades ago. The ‘yield’ was supposed to be highest at this time point in pregnancy, for which metabolic-endocrine explanations were suggested. ^*[20,21]*^ However, majority of women diagnosed with GDM had elevated glucose in early pregnancy even in an American clinic, suggesting a substantial contribution of pregestational glucose intolerance. ^*[22]*^ It is noteworthy that a comparable prevalence of glucose intolerance in young non-pregnant American women to that in pregnant women, led to a suggestion in 1988 that GDM could represent undiagnosed pre-gestational hyperglycemia. ^*[23]*^

A substantial range of risk factors for GDM (obesity, central obesity, dysglycemia, dyslipidemia, NAFLD, chronic inflammation, PCOD and other endocrine disturbances) are present pre-gestationally [3, 4], and track from childhood into adult age. ^*[24]*^ Thus most of the risk factors of GDM are already present before conception and expose the fetus to adverse conditions in the peri-conceptional period. Frequent presence of a large fetus, polyhydramnios and higher abdominal wall obesity in the fetus on ultrasound examination at the time of diagnosis of GDM suggest ongoing hyperglycemia from early pregnancy. ^*[25]*^ Higher prevalence of NTDs and other congenital anomalies in GDM also suggests peri-conceptional metabolic disturbance. ^*[26,27]*^ The pregnancy related factors which increase risk of glucose intolerance relate to gestational weight gain, multiple pregnancy and male fetus in addition to the endocrine and other influences from the placenta. ^*[3, 4, 28]*^

Research and clinical experience suggest that peri-conceptional period may be the most crucial in fetal programming of NCDs and there is a growing consensus that improvement in maternal nutrition and metabolism starting from before conception is important to influence the risk of NCDs in the offspring. ^*[10, 29,30]*^ Our data suggests that the so called ‘GDM’ may partly represent undiagnosed, pregestational hyperglycemia. Role of pre-conceptional improvement of maternal glucose to prevent fetal anomalies in diabetic women is well established ^*[31]*^, and it is interesting that a pre-conceptional supplement with micronutrient rich snack reduced the incidence of ‘GDM’ in the Mumbai slums. ^*[32]*^

Our findings also raise the possibility that the failure of GDM treatment to reduce risk of childhood obesity and glucose intolerance in the offspring in the randomized controlled trials may be related to a contribution of peri-conceptional programming because the women were investigated and treated for GDM only in the third trimester. ^*[33,34]*^ Our findings also reinforce the need for intensifying the clinical protocol of post-partum follow up of GDM women to diagnose and treat glucose intolerance and to institute preventive measures even in those who become ‘normoglycemic’ after delivery.

Our paper has many strengths and some weaknesses. This is one of the few studies to evaluate glucose tolerance of GDM women in India within one year of delivery. Even though the screening for GDM was based on ‘high risk’ approach at the time of the study, the participants are representative of those in many of the hospital based diabetes clinics in Indian cities. KEM hospital cares for a wide section of the population and therefore, there is a fair representation of the lower and upper middle class women. The criteria for diagnosis of GDM (plasma glucose concentration >140 mg% 2 h after 75g oral glucose load) are similar to the widely used DIPSI criteria in India. Even though the participation rate was ∼ 50% for the postpartum follow up, there was no difference in the levels of risk factors for diabetes between those who attended and those who did not, reducing the chances of bias in our findings. Inclusion of matched non-GDM comparator group allows interpretation of findings in relation to the background prevalence of glucose intolerance in this population. All these factors help to make our study relevant for the clinicians as well as the policy makers to consider introducing pre-pregnancy assessment of glucose tolerance in young women, at least in the high-risk group.

We suggest that the so-called ‘GDM’ in the high risk developing populations of the world may represent undiagnosed pregestational diabetes in a substantial proportion of cases. This possibility needs to be tested in a wider population in different countries to revise the policy for timing of screening for ‘pregnancy’ hyperglycemia.

## Data Availability

Data is available on request with Dr CS Yajnik at

## Acknowledgement

To the staff of Diabetes Unit and participants.

